# Determining factors for pertussis vaccination policy: A study in 5 EU countries

**DOI:** 10.1101/2019.12.11.19014449

**Authors:** Anabelle Wong, Annick Opinel, Simon Jean-Baptiste Combes, Julie Toubiana, Sylvain Brisse

## Abstract

Pertussis vaccination policy varies across Europe, not only in the type of vaccine – whole cell (wP) vs. acellular (aP1/2/3/5) – but also in the schedule and recommendation for parents. This study aims to investigate the determining factors for the type of vaccine, immunization schedule and maternal immunization recommendation. From March to May 2019, experts in national health agencies and major academic or research institutions from Denmark, France, Poland, Sweden and the UK were invited to a semi-structured interview. Thematic analysis was performed on the transcripts using a codebook formulated by 3 coders. Inter-coder agreement was assessed. Fifteen expert interviews were conducted. The identified driving factors for pertussis vaccine policy were classified into three domains: scientific factors, sociological factors, and pragmatic factors. The determining factors for the type of vaccine were prescriber’s preference, concern of adverse events following immunization (AEFI), effectiveness, and consideration of other vaccine components in combined vaccines. The determining factors for infant schedule were immunity response and the potential to improve coverage and timeliness. The determining factors for maternal immunization were infant mortality and public acceptability. To conclude, socio-political and pragmatic factors were, besides scientific factors, important in determining the vaccine type, schedule of childhood immunization and recommendations for parents.

## 1. Introduction

Whooping cough is an acute respiratory infection characterized by repeated, intense cough bouts that can last for 2 to 3 months [1]. In 1906, the pathogen causing whopping cough was found to be a Gram-negative bacterium, *Bordetella pertussis* (*B. pertussis*) [2]. It is highly contagious and spreads via droplets of the coughs or sneezes of an infected person [1]. Before vaccines became available, it was a major cause of infant mortality [3].

The earliest vaccines against pertussis were inactivated whole cell pertussis vaccines (wP) developed in 1930s to 1950s; routine pertussis vaccination in children less than 12 months started in the late 1950s in Europe [4]. wP have been replaced by acellular pertussis vaccines (aP) in many European countries since the 1990s [2,5]. aP do not contain the whole bacterium but only antigens; it was suggested that aP have lower reactogenicity and are better accepted [3].

After more than half a century, whooping cough has yet to be eradicated and there are signs for resurgence despite routine childhood immunization [3,6] with high coverage [5]. Some authors have hypothesized that the resurgence is due to short-lived protection from aP compared to wP [6,7]. Nevertheless, the waning of immunity from aP cannot explain all of the resurgence. For example, in the Netherlands and in Denmark, the incidence of whooping cough began to surge before wP were replaced by aP in the national immunization program [6,8]. Evidence from recent research supports the adaptation of *B. pertussis* to vaccine-induced immunity through antigen evolution [7,9]. Most notably, some strains of pertussis no longer produce pertactin (PRN) [3,4,5,10], a protein that enables the bacteria to attach to the lining of human’s airway [11] and which is one of the components used in 3-component (aP3) and 5-component (aP5) vaccines [3,4,5,10,11]. The PRN-negative characteristic appears to give *B. pertussis* an advantage in surviving in aP-vaccinated populations [5,11,12,13]. However, other vaccine components continue to provide protection against *B. pertussis*. Currently, there is no evidence that the PRN-negative strains cause more severe pertussis infection [14,15] and one study showed that the proportion of apnea was lower among PRN-negative cases [12].

Pertussis vaccination is an indispensable strategy towards disease control and prevention. However, the dynamics of vaccination policy are complex. Understanding determinants of vaccine policy is a step forward in the optimization of national immunization efforts. The scope of this study focuses on the relationships of the content and context of pertussis vaccination in Europe. The content is defined as i) the type of vaccine, ii) national immunization schedule and iii) recommendations for pregnant women in pertussis vaccination; while the context comprises determining factors for the policy.

The type and schedule used in the national immunization programs in Europe have evolved since the introduction of pertussis vaccine in 1950s (Appendix A) [3,4,6,16-50], leading to considerable heterogeneity in pertussis vaccination policies across Europe. Currently, the most common type of vaccine used in Europe is multi-component aP (aP2/3/5). Two-component aP (aP2) contains two antigens: pertussis toxoid (PT) and filamentous hemagglutinin (FHA); aP3 contains the additional antigen PRN; and aP5 contains PT, FHA, PRN, and fimbriae (Fim) types 2 and 3 [4]. While France allows the use of all three types of multicomponent aP, Denmark had until recently relied exclusively on a 1-component aP vaccine (aP1) that contains only PT [51]; whereas Poland remains the only country in Europe that uses wP in the national childhood immunization scheme [46]. Given the existing evidence on efficacy and safety of pertussis vaccines and the similar profiles of *B. pertussis* strains circulating in Europe [3,4,5,9,10], such diverse vaccination policies across Europe lead to the hypothesis that pertussis immunization strategy is not solely determined by scientific factors but may also be influenced by socio-historical factors as well as pragmatic reasons.

Two main patterns of the first series of pertussis immunization schedules for children are currently used in Europe [16]: i) the accelerated schedule: 2/3/4 or 2/4/6 month with or without the 4th dose before the age of 2 years; and ii) the long schedule: 2/4/11 or 3/5/12 month.

The accelerated schedule consists of 3 doses of vaccines in the first 6 months of life whereas the 3 doses of vaccines in the long schedule are given in a span of 11 to 12 months [52]. The immunization schedules vary among countries using the same type of vaccine. The initiation of infant immunization can be at 2 or 3 months. Studies conducted in the 1990s did not offer conclusive evidence. Some studies suggested higher serological response using the long schedule [53,54]; however, no good serological correlate of protection has been identified [52]. Systematic review also found no good data for the comparison of different schedules in terms of effectiveness [52]. Besides, the evidence of the age of infant immunization initiation having an impact on immunogenicity or effectiveness is limited [52]. Under such circumstances, the observation of the variation of infant immunization schedule further supports the hypothesis that pertussis immunization strategy is influenced by factors other than existing evidence in efficacy or effectiveness. This study aimed to investigate the factors that determined the pertussis immunization strategy in European countries that have distinctive vaccination policies.

## 2. Materials and Methods

### 2.1. Selesction of countries

Official reports from national health agencies and scientific and medical journals were reviewed [3,4,6,16-50] to provide information on the current pertussis vaccination policy in 11 EU countries that have participated in any one of the 4 EuPertStrain studies (Appendix A). The EuPertStrain studies were established within the European network in 2011, aiming to monitor changes in the European *B. pertussis* populations in order to optimize vaccine strategies [3,4,5,10]. Figure 1 shows the type of vaccine used and the first series schedule of childhood immunization in 11 EU countries. Five of these countries were then selected for a qualitative study based on the type of vaccine being used, different recommendations for parents, and different schedules for childhood immunization.

**Figure 1.**
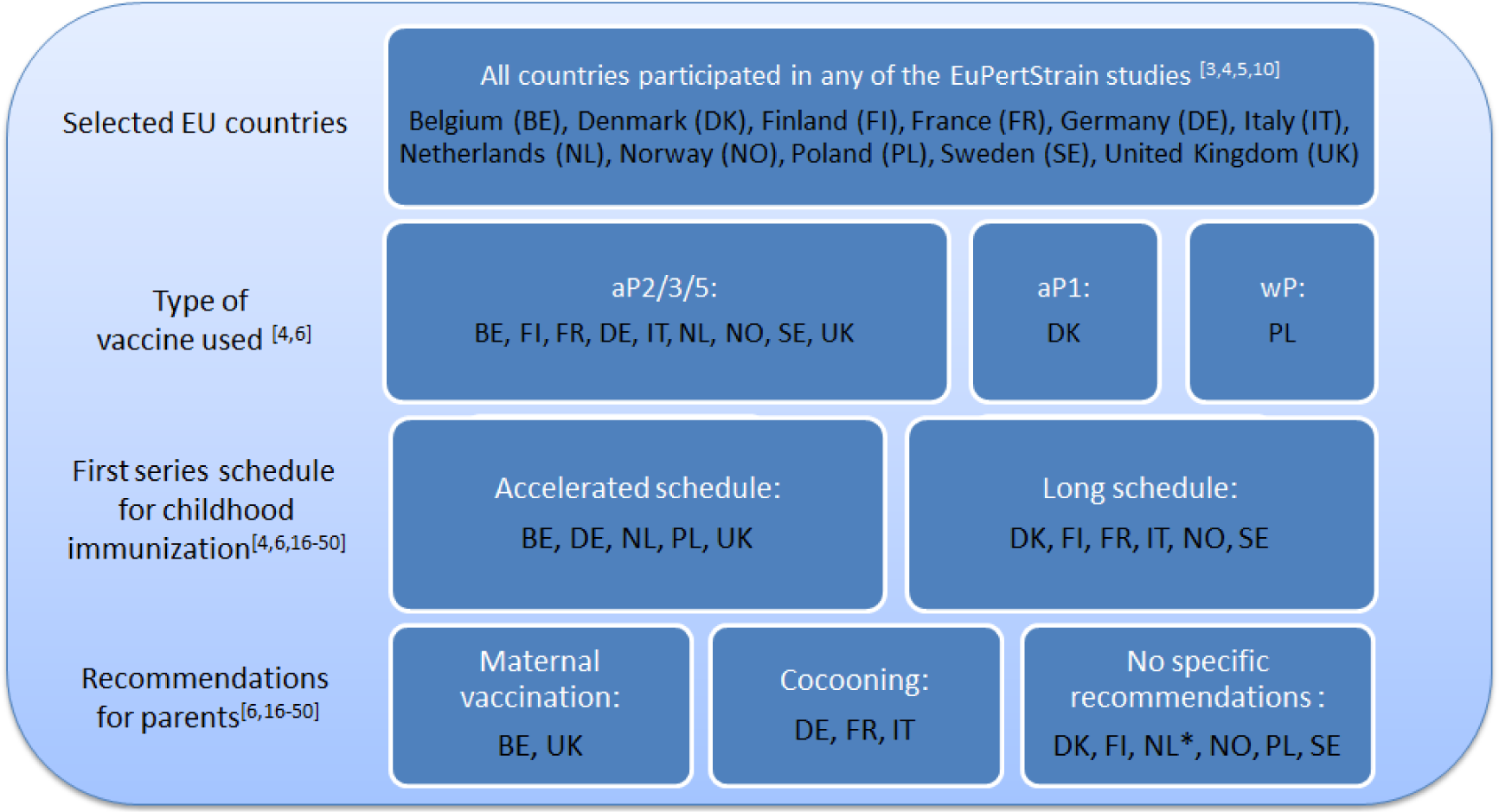
The type of vaccine used, the first series immunization schedule and recommendations for parents in 11 EU countries (*NL started recommendations for maternal vaccination during the study).

Denmark and Poland were selected due to the unique immunization agents used. The UK was selected as it is the first country in Europe to have implemented mass vaccination for pregnant women [1]. France was selected due to its recommendation for the cocooning strategy – a strategy that aims to fill the vulnerable gap for pertussis infection in children between 0 to 6 weeks by vaccinating the individuals in close contact with the newborn [16,33]. Owing to a unique history of using aP1 exclusively in Gothenburg from 1996 to 1999 [50] and its “vaccine vacuum” from 1979 to 1996, Sweden was also included in this study to contribute information about vaccine policy evolution. In terms of childhood immunization schedule, Poland and the UK adopted the accelerated schedule while Denmark, France and Sweden followed the long schedule.

### 2.2. Methods for interviews

Semi-structured interviews with experts in selected countries were conducted to gain understanding on the factors determining pertussis vaccination policy. Key informants were selected due to their role, experience and knowledge in the field of childhood immunization [55]. The purpose of a semi-structured interview was to encourage expert participants to share their observation and understanding about the phenomenon in vaccination policy making. Experts were identified in the process of literature review and via searching the official websites of major academic and national institutions in selected countries. Individuals who fulfilled the inclusion criteria (Table 1) were recruited. Further informants were recruited by snowball sampling because sampling in this study was purposive and it aimed at maximum variation of information [56]. The sampling frame aimed to recruit experts from different professional backgrounds within the same country and to obtain representation from experts involved in policy decisions as well as those who were not involved in the national decision process. Interviews were conducted in person or via Skype or phone when a face-to-face meeting could not be arranged. The interviews followed a set of open-ended questions in the topic guide developed based on Boyce and Neale’s template [57]. The interviews were audio-recorded and transcripts were produced based on the audio-file. The transcripts were sent back to participants for checking and signing as an endorsement of accuracy. Upon receiving the endorsed transcript, the audio-recording would be deleted. In cases where the expert had not sent back an endorsed transcript, draft transcript was used for analysis while pending future transcript endorsement to trigger the deletion of the audio-file. The transcripts were anonymized by the allocation of a transcript ID.

**Table 1.** Inclusion and exclusion criteria for participant recruitment.

| Inclusion Criteria |
| --- |
| 1) Have work experience of 3 years or above in the field related to vaccination policy or pertussis research in the country; |
| 2) Have professional knowledge on pertussis vaccine or vaccination policy in the country; |
| 3) Are able to communicate effectively in English; |
| 4) Are willing to participate and have given informed consent. |
| Exclusion Criteria |
| 1) Vaccine manufacturers and their employees; |
| 2) Withdrawn consent during or after interview. |

Demographic information and characteristics of participants were summarized by descriptive statistics. Thematic content analysis was performed on the anonymized transcripts using a codebook formulated based on grounded theory. It means that the codebook did not rely on any pre-existing theory but consisted of recurrent concepts emerged from the interviews; therefore the codebook was considered grounded in data [58]. The process of coding was guided by the 3-stage approach proposed by Campbell and colleagues [59]. In the first stage, a codebook by one knowledgeable coder was developed based on all transcripts. A knowledgeable coder was defined as a coder who had insights on all transcripts. This codebook was used by three independent analysts to code two pilot transcripts: one from the interview with a health scientist and the other with a social scientist. As the vocabulary used and the concepts brought up by experts with these professional backgrounds can vary significantly, the code used would vary accordingly. The second stage involved adjudicating coding disagreement through discussion [59]. In this stage, the codebook was refined as ambiguous and overlapping codes were deleted and necessary new codes created based on the consensus of the three coders. Inter-coder agreement was assessed after pilot coding. In the third stage, the codebook was deployed to the full set of anonymized transcripts by one knowledgeable coder once inter-coder agreement was established. All codes were arranged into categories and plotted for conceptualization. All transcripts were processed in R version 3.5.1 and RStudio version 1.1.456 using the package “RQDA”.

Inter-coder agreement was assessed using Krippendorff’s alpha, which is a generalized version of inter-rater agreeability statistic that can be applicable in nominal data and in situations where there are more than 2 observers [60]. The interpretation of Krippendorff’s alpha was based on the guideline drawn up by Landis and Koch [61].

### 2.3. Ethicalapproval

The study has obtained approval from the Ethical Committee (EC) of the University of Sheffield, UK. Regarding the nature and study design of this study, EC approval or notification was not required in Denmark, France, Poland and Sweden. Further to EC requirements in selected countries, this study observed and complied with the EU General Data Protection Regulation (GDPR) as well as the country-specific regulations if the scope of such regulation applied.

## 3. Results

From March to May 2019, 34 experts were contacted, and 15 interviews had been conducted by the end of May, 2019. The 15 experts had experience ranging from 5 to 35 years and the median year of experience was 18 years. Table 2 shows more demographic information including country, professional background and whether the expert was involved in the vaccination policy process.

**Table 2.**
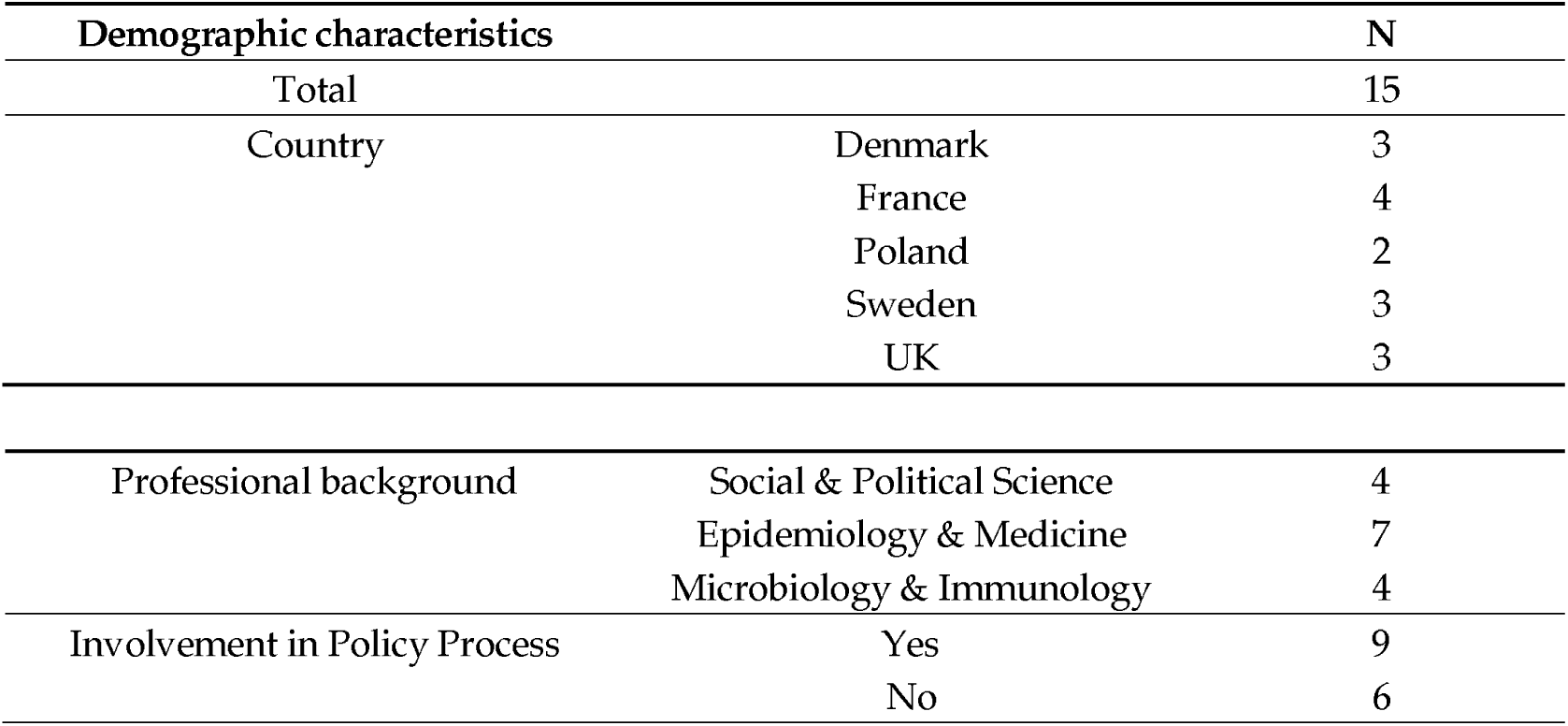
Demographic characteristics of participants.

Among the 19 non-responders, 6 declined for not having sufficient knowledge or experience in the topic, not being actively involved in related duties anymore, or being too occupied with other priorities. The characteristics of the non-responders were analyzed. Non-responders more often came from Sweden and the UK (n=15); and were more often experts in the field of social and political science (n=10).

### 3.1. Inter-coder agreement

Inter-coder agreement among the three coders was assessed using Krippendorff’s alpha (Table 3). Disagreements among coders came from the inconsistency within one coder as well as the inter-coder differences in the interpretation and application of the coding guidelines [60].

**Table 3.** Inter-coder agreement in pilot coding using two transcripts.

| Transcript Profile | Coding Unit | Krippendorff's alpha<sup>[60]</sup> | Interpretation<sup>[61]</sup> |
| --- | --- | --- | --- |
| Epidemiology & Medicine | 75 | 0.621 | Substantial |
| Social & Political Science | 57 | 0.598 | Moderate |
| Total | 132 | 0.616 | Substantial |

The Krippendorff’s alpha based on the health scientist’s transcript was slightly higher than that based on the social scientist’s transcript. The small difference between the Krippendorff’s alphas of the 3 coders based on the 2 transcripts also suggested that the levels of agreement among coders were similar when interpreting discussions offered by experts of different backgrounds. The overall inter-coder agreement by Krippendorff’s alpha was 0.616, indicating substantial agreement according to Landis and Kock’s interpretation framework (Table 2) [61].

### 3.2. Determining factors of pertussis vaccination policy

The determining factors referred to the reasons for the change in policy or the ground for policies remaining unchanged. These factors were identified from the interviews and triangulated by experts from the same country.

The codes derived from the transcripts were categorized into three domains: scientific factors, sociological factors, and pragmatic factors. Figure 2 shows the determining factors for pertussis vaccination policy under different domains.

**Figure 2.**
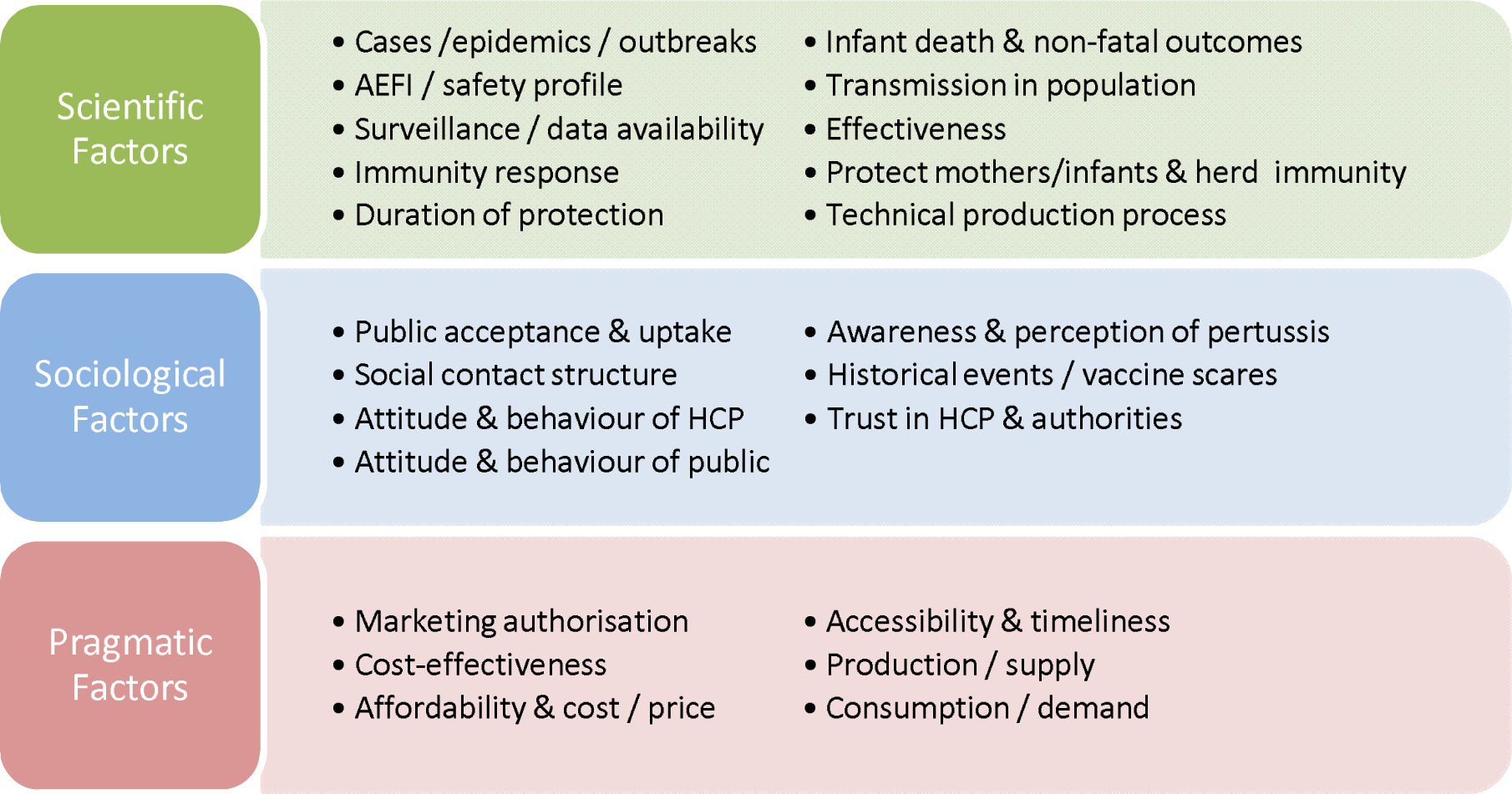
Determining factors of pertussis vaccination policy: (**a**) scientific factors; (**b**) sociological factors; (**c**) pragmatic factors.

Scientific Factors

#### 3.3.1. Type of vaccine

Regarding the change from wP to aP, a range of reasons were mentioned. While most experts (n=10) believed the concern of adverse events following immunization (AEFI) was causing the change, some experts who were involved in the decision making process pointed out other more proximal reasons (n=5) for the change in the type of vaccine being used in the national immunization program.

Experts in France stated that the prescriber’s preference contributed to the change. According to the experts, the country recommended using wP in the first series of childhood immunization until 2004; however, since the introduction of aP booster for the age group of 11 to 14 years in 1998 in France and subsequent availability of aP, many general practitioners and pediatricians prescribed aP for the prime vaccination of the infants. According to the experts, due to increasing consumption of aP and decreasing demand of wP, manufacturers decided to produce only aP instead of maintaining two production lines. That ultimately caused the change in recommendation from using wP to aP in the immunization for all age groups.

Experts in Sweden pointed out the fact that despite a high coverage of wP before the change, pertussis cases in infants were not reduced. Therefore, the wP vaccine was deemed ineffective and the national pertussis immunization program was suspended from 1979 to 1996. It was resumed when aP became available and after its effectiveness had been proven.

In the UK, wP was recommended for the infant immunization program until 2004. According to the experts, coverage of the vaccine program plummeted due to concern about AEFI in the 1970s and 1980s, but confidence was restored in wP given abundant efforts in independent review about the safety of the wP vaccine, leading to increased and sustained high coverage. The change from wP to aP was actually driven by the need of an inactivated polio vaccine, which was available in a multivalent vaccine that contained an aP component.

Poland was the only country in the EU that is still using wP. According to the experts in Poland, the wP used had desirable effectiveness and did not induce concern about AEFI. As the vaccine was produced in the country, the supply was stable. It was also a more affordable option.

Regarding the switch from aP1 to aP2/3/5, experts from Sweden shared that it was the tender process that drove the change. As aP1 vaccines were only produced in Denmark, since the manufacturer did not participate in the tender process, it was not available for selection in the Swedish national immunizations program. Before this study commenced, Denmark was the only country that had been using aP1 [51] since the change from wP to aP in 1997. However, experts in Denmark reported in the interview during this study that the country has just changed from using aP1 to aP2/3/5 and the reason was also due to aP1 manufacturer stopping its participation in the tender process.

#### 3.3.2. Immunization schedule

Concerning the accelerated and long schedule for infant immunization, most experts regarded it to be a decision based on clinical data from trials of the vaccines and the authorized posology recommended by manufacturers. However, since both schedules were proven to be effective and the recommendation by the World Health Organization (WHO) also allows certain flexibility [62], countries made their decisions based on various factors and priorities.

Experts from Denmark and Sweden, where long schedule (3/5/12 month) was used, shared that such decision was based on data from clinical trials within the country and that such schedule allowed desirable immunogenicity and effectiveness.

> *“That was in the 80s, we looked at the pertussis trial and we found that the immune response is the best if you give the vaccine at 2-month intervals instead of 1-month intervals. And it’s better to start at the age of 3 month compared to 2 month…”* (Sweden expert)

France adopted the accelerated schedule in 1995 but has changed to a long schedule in 2013.

However, in 2013, France used a long schedule that consisted of three doses at 2, 4, 11 month instead of the 3/5/12 month standard long schedule. According to the experts, an earlier initiation of immunization was due to the concern of cases in very young infants – those who might be infected by pertussis before receiving the first dose of vaccine at the age of 3 month according to the standard long schedule.

> *“And in France, we decided to adopt such a schedule but we did not want to start at 3 month because we knew that if you start one month later, you will have more pertussis cases. So we chose to start at 2 months…”* (France expert)

The accelerated schedule (2/3/4 month) was used in the UK. Experts shared that when the 3rd dose of primary pertussis vaccination was closer to an older infant age around 11 month; there was a drop-off in uptake of that 3rd dose. By adopting the accelerated schedule, coverage was increased and timeliness of the three doses of first series of vaccine was improved.

> *“The advantage of having an accelerated course is that you’re not only offering protection at an earliest infant age but also we’ve found that you’re more likely to achieve higher uptake, when you’re offering it at an earlier age*.*”* (UK expert)

#### 3.3.3. Maternal vaccination

A consensus was observed among experts from different countries that the main reason for recommending vaccination during pregnancy was an epidemic or increased infant death.

> *“…we have no, well, very few infant deaths. And that is the major marker for introducing the vaccination in pregnancy, of course. We had an epidemic in 2016. And that’s prompted, the Health Authority to think about renewing the vaccination strategies. And vaccination in pregnancy was one of them although we haven’t implemented it*.*”* (Denmark expert)

> *“So now it has changed in favor of maternal vaccination. Pertussis is not a big problem for the moment, but in case there will be an increase of infants infectedwith pertussis, I think there will be a change in the program towards this recommendation*.*”* (Sweden expert)

> *“We experienced a very significant increase in overall rate of disease across the entire population, but particularly in those very young babies. And we had an increase in pertussis deaths, we had 14 deaths from pertussis in 2012. So the introduction of the maternal program was very much introduced and prompted by the increased rate of disease. We’ve done it as an emergency program…”* (UK expert)

In countries where infant deaths remained low, such as Denmark and Sweden, experts suggested that maternal vaccination would enter the policy discussion when pertussis-related infant deaths increased. In the UK, where a major epidemic with increased infant deaths occurred in 2012, such strategy was adopted as a response to the emergency.

However, there were also recurrent concerns about such policy. These concerns included acceptability of maternal immunization by health care professional (HCP) and by the general public and the limited data on immunity blunting, which is a phenomenon where trans-placentally acquired antibody lowers the immune response to infant immunization [19].

> *“Well, it’s typical that one reason why we didn’t jump at that strategy is that we have the question mark of what will be the acceptance rate of such a strategy*.*”* (France expert)

> *“…we have an investigation in the public health agency a couple of years ago. At that time, there was some hesitancy. They were looking for more data. There’s some kind of blunting or immune response in the children*.*”* (Sweden expert)

#### 3.3.4. Other important discourse: policy implementation

During the interviews, some experts (n=2) expressed that pertussis vaccination was not a frequently debated topic in the country as vaccination debates focused on other vaccines, such as HPV vaccines.

Some experts (n=3) offered a discussion on the paradigm of vaccination policy process. In Poland, there was recently a vote in parliament about abolishing mandatory vaccination, which was triggered by citizens’ petition; and in France, there had been a citizen consultation before the country expanded mandatory vaccination in 2018 from 3 vaccines to 11 vaccines. Those events led countries to discuss themes related to public-professional and public-authority relationship. Some experts shared that they were concerned about how policy was made would have an impact on the perception, attitude or behavior of the general public regarding vaccination or towards HCP:

> *“And something that might fuel distrust towards the expert professionals, in Poland at least, is the strength and unquestionability of the consensus already existing among professionals*.*”* (Poland expert)
>
> The use of mandatory vaccination may lead some people to be more radical:

> *“…*.*that doesn’t mean they were against vaccines, it just means that they were against mandatory vaccination… is it going to push part of the people who are hesitant towards a more radical stance, you know, going to very private schools, where they look the other way and don’t really check whether the children are vaccinated? And the issue is whether it’s going to create small pockets of severely under-vaccinated people…”* (France expert)
>
> According to the some experts, mandatory vaccination may be seen as a way to restore public confidence in vaccines in France and as the consensus or approval of the authority and professionals in Poland. Other countries found voluntary vaccination based on national recommendation a better suiting strategy. Some experts also expressed doubts about the message or implication conveyed by vaccine mandates:

> *“I can’t see a reason for introducing a compulsory element to this because it’s a program that’s already very well delivered and very well received. So I can’t see a role for mandating in our population at this time. And I doubt it would improve uptake, and it could be counterproductive*.*”* (UK expert)

> *“…but if the results speak for themselves and the health authorities recommend something, people tend to do that. And I believe, and I know I’m not alone in believing that, if we were to make mandatory vaccination, it would actually spark this hesitancy, it would spark distrust. And I think it would be detrimental for our program to do that*.*”* (Denmark expert)

## 4. Discussion

In this work we first reviewed the literature to define the extent of variation of vaccination policies across EU countries. We then selected 5 countries with contrasted policies to conduct an investigation of vaccination policy determinants. In the semi-structured interview, experts from Denmark, France, Poland, Sweden and the UK were asked about their perception of the determinants for changes in the type of *pertussis* vaccine (wP / aP1 / aP2,3,5) and schedule being used in the national childhood immunization program, and the reasons for such changes, if any. Experts were also asked why cocooning strategy or maternal vaccination might be important to the country, if there was such recommendation in place.

All participating countries except Poland experienced the switch of wP to aP in national immunization against *pertussis*. Although it is widely believed that aP is better tolerated than wP [3], it was not the main reason for the switch in France, Sweden and the UK. Moreover, the wP in Poland did not trigger concern about AEFI. An explanation for these observations is that wP, of which the production process is not standardized, showed different efficacies and safety profiles in different countries, leading to heterogeneity in the discontinuation or continuation of wP in national immunizations program.

One of the recurrent messages brought up by experts was the inappropriate comparison between wP and aP in literature and in nowadays’ debates. Experts reminded researchers and policy makers that wP and aP comparison would not be meaningful and can be misleading when the strain of wP was not specified or characterized. The efficacy and safety profile of wP depended on the specific strain and the production of the vaccine. Some countries used locally produced wP; therefore the quality, efficacy and safety profile of the vaccines across countries would vary. Even within the same country, wP still varied from year to year, and from batch to batch. This also explained why the decision of using wP or aP and the reasons for the switch differed from country to country. If the country had a wP with good effectiveness and desirable safety profile, wP tended to remain in use for longer periods. As different wP have different safety profiles, it is therefore important to be precise about the wP when the concern of AEFI is addressed.

Often mentioned was the “side effects” or “safety profile” of wP but the more important concept is the distinction between the actual AEFI that occurred and the concern about the AEFI. The former is a factual concept – frequency and severity of the AEFI that occurred; the latter is a concept about perceived risk and a measure of attitude.

Looking closer at the driving factors for the change from wP to aP in the selected countries, there appeared to be an interdependent relationships among scientific factors, sociological factors and pragmatic factors. This study leads us to propose a mechanism of influences among cases of AEFI, the concern about AEFI and the behavioral adaptation (Figure 3). Hidden influence or relationship, such as the influence from manufacturer on HCP and policy makers, or the influence from media and social media on HCP and policy makers, can be present. The existing literature on vaccination policy also highlighted that transparency of the decision process needs improving [63]. Therefore, while deciphering the driving factors in vaccination policy, one must bear in mind the potential hidden driving factors or the concealed relationship between factors.

**Figure 3.**
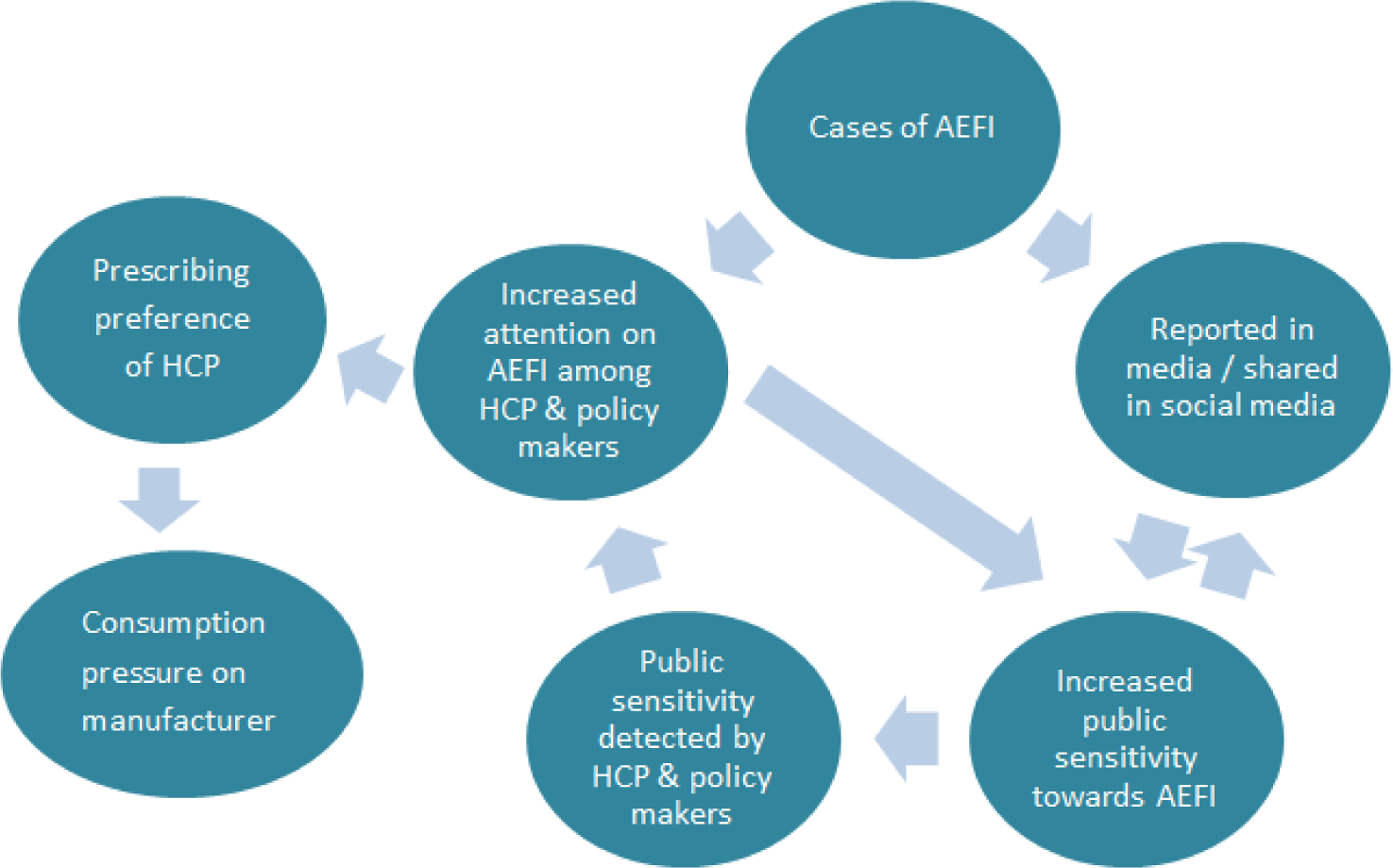
Relationship between AEFI and the concern about AEFI among public and HCP.

The determination of the childhood immunization schedule against pertussis appeared to result from a balance of three essential factors: i) immunogenicity; ii) earliest immunization possible; and iii) highest uptake possible. These three independent but important factors justify the flexibility included in the WHO recommendations [62] and offer a basis for vaccine policy makers to weigh the above-mentioned factors according to the country-specific context.

Closely related to the determination of the childhood immunization schedule are the recommendations for parents. Most experts agreed that maternal vaccination became a topic of discussion when an epidemic or an increase in pertussis-related infant deaths was observed in the country. Such finding is coherent with the policy evolution in the UK, as the UK government introduced a temporary immunization program for pregnant women from October 2012 as a response to the national outbreak declared in April 2012 [64]. Later in June 2014, the Joint Committee on Vaccination and Immunisation (JCVI) advised the program to continue for another five years before further evaluation [64]. Such evaluation shall offer an abundance of evidence that may answer many questions about the longer term safety, efficacy and cost-effectiveness of the national maternal immunization program.

A similar case was observed in an oversea Department of France: the increase in pertussis cases among infants in Mayotte in 2017 has sparked the discussion that led to the implementation of local maternal immunization as a response to the epidemic [65]. The outbreak in Mayotte was partly due to insufficient coverage of childhood immunization program caused by the breakdown of health care infrastructure [65]. Further, the Technical Commission of Vaccinations (Commission Technique des Vaccinations, Haute Autorité de Santé) has affirmed that maternal vaccination is more efficient and beneficial than the cocooning strategy in protecting infants too young to be vaccinated in the context of major epidemics [66]. Hence, the situation in Mayotte might influence an evolution of maternal vaccination recommendations even in mainland France.

As primary research, this study offered abundant data and will allow further analyses. Its sampling frame aimed to obtain representation from all five selected countries. In each country, at least two different professional backgrounds were included in order to maximize data variation and to serve the purpose of triangulation. The median duration of experts’ experience was 18 years, indicating that the study sample has long duration of exposure in the field of interest and can be considered as a credible source of information. The data collection relied on audio-recording, together with the transcript endorsement process, which ensured the accuracy of data and thus increased the internal validity of the study. Qualitative research is often attacked for being subjective and biased [67], especially when there is ambiguity in the data or when certain codes belong to more than one coding categories [68]. To increase rigorousness and external validity, this study endeavored to establish a reliable codebook with an assessment of the inter-coder agreement of three coders who had worked on the same set of transcripts [59,60]. Although the sample size was small (n=15), no new code arose towards the last few interviews and a large amount of recurrent codes were observed, suggesting saturation of knowledge, which was the purpose of this qualitative research using the method of interview [69].

There were a few limitations in this study. Firstly, the study sample included more scientists from the natural and health disciplines than social and political ones, which caused the data to be more science-centric. The reason for such sample composition was driven by the reality that more experts from the field of natural science and health science responded to the study invitation than experts from the field of social and political science. This could be due to no or very few sociologists and political scientists working on pertussis vaccination specifically, and some may not feel comfortable participating in a study on pertussis vaccination policy. Secondly, the set duration of 30 minutes was a limiting time frame for an interview, so some participants did not cover all topics on the interview topic guide. Thirdly, as discussed above, hidden external influence such as that from the pharmaceutical industry cannot be assessed. This may or may not have affected the neutrality of experts’ opinions [70]. Since participants did not declare conflict of interests, it was not possible to estimate the impact of such influence on this study. As with many studies using the method of qualitative interview, unitization of transcripts was a limitation in this study [59,60]. In the content analysis, coding units were “units of meaning” instead of demarcated parts of text. Such unitization, despite being more appropriate in this exploratory research study using complex interview data [59], tends to result in lower inter-coder agreement [59,60]. Lastly, the five countries had different surveillance systems and strategies in place, which may influence the experts’ perception of determining factors for pertussis vaccination policy.

## 5. Conclusions

By disputing the oversimplified version of safety- and efficacy-driven vaccine policy, the findings of this study contribute to a better understanding of the determining factors that drive pertussis vaccination policy. The choice of the immunization agent was influenced by prescriber’s preference, concern of adverse events following immunization (AEFI), effectiveness, and consideration of other vaccine components in combined vaccines. The schedule of childhood immunization was determined by immunity response and the potential to improve coverage and timeliness. The recommendations on maternal immunization hinges on infant mortality contributed by pertussis in the country as well as acceptability of such strategy by HCP and the general public.

To better guide pertussis vaccination policy, future researchers should pay attention to should pay attention to the impact of changes in vaccine policies on pertussis epidemiology within a country and dynamics of transmission in Europe and worldwide. More efforts and attention should be given to sociological research on pertussis vaccination strategies, especially on the attitude and behavior of HCP and the public. Including sociological expertise in the decision making and increasing the transparency of decision process may also help build public’s trust in the vaccine policy decision process.

## Data Availability

All data are available and archived; transcripts details are not to be published due to confidentiality agreement.

## Author Contributions

Conceptualization, Anabelle Wong, Annick Opinel, Julie Toubiana and Sylvain Brisse; methodology, Anabelle Wong and Annick Opinel; software, Anabelle Wong and Simon Jean-Baptiste Combes; validation, Anabelle Wong, Annick Opinel and Simon Jean-Baptiste Combes; formal analysis, Anabelle Wong; investigation, Anabelle Wong; resources, Annick Opinel, Julie Toubiana and Sylvain Brisse; data curation, Anabelle Wong; writing—original draft preparation, Anabelle Wong; writing—review and editing, Annick Opinel, Simon Jean-Baptiste Combes, Julie Toubiana and Sylvain Brisse; visualization, Anabelle Wong; supervision, Annick Opinel and Simon Jean-Baptiste Combes; project administration, Anabelle Wong and Annick Opinel; funding acquisition, Sylvain Brisse.

## Acknowledgments

The research team would like to thank Yoriko Masunaga, Institute of Tropical Medicinein Antwerp; Richard Cooper, University of Sheffield; Didier Guillemot, Tamara Giles-Vernick and Matthieu Domenech de Cellès, Institut Pasteur, for their valuable advice and resourcefulness. We would also like to express our heart-felt gratitude to Daniel Levy-Bruhl, Daniel Floret, Nicole Guiso, Tine Dalby, Michal Zabdyr-Jamróz, Sven-Arne Silfverdal and other experts who have offered their support to this project.

## Appendix A Pertussis Vaccination Policy in 14 EU Countries since Introduction. [3,4,6,16-50]

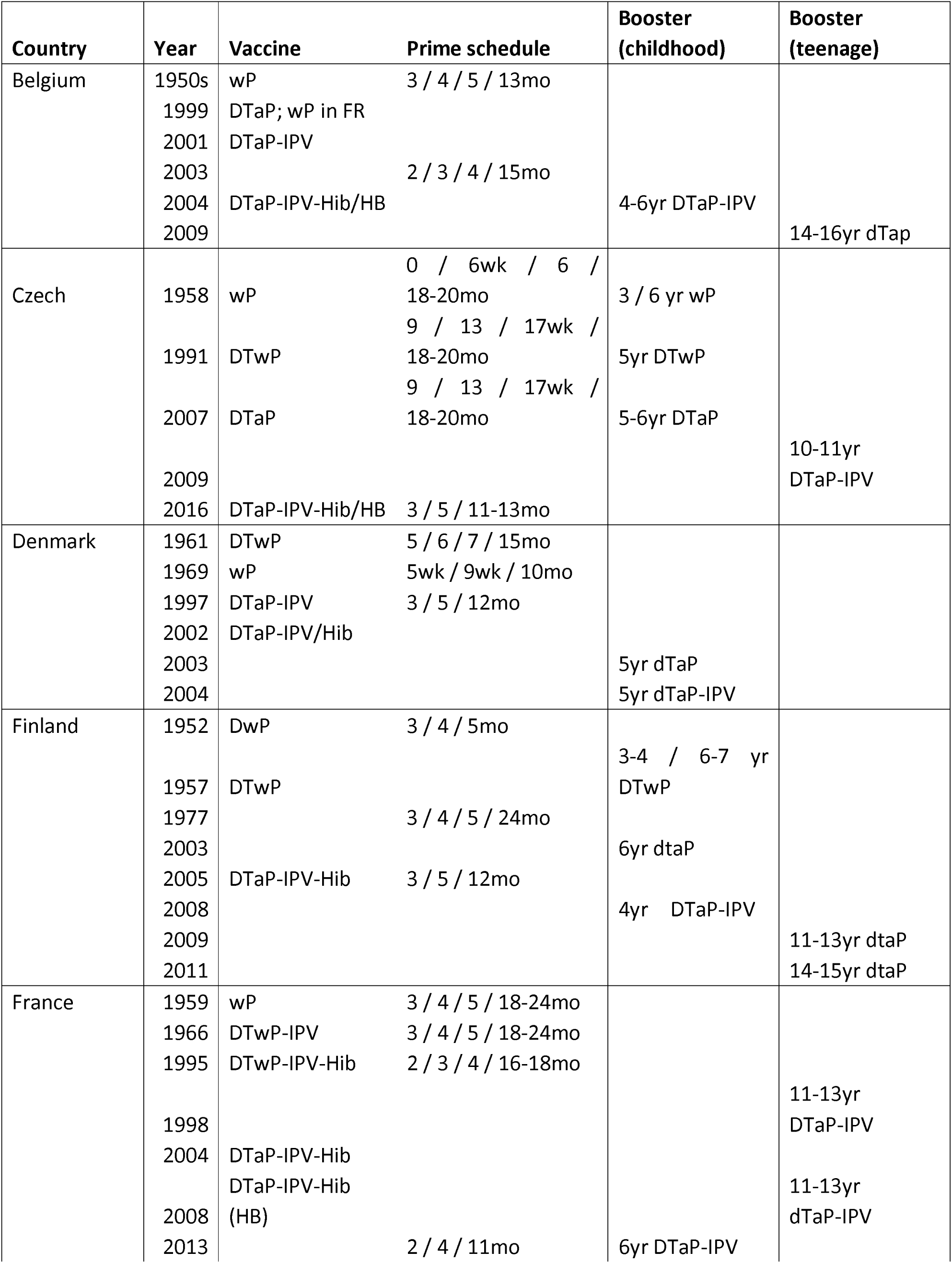
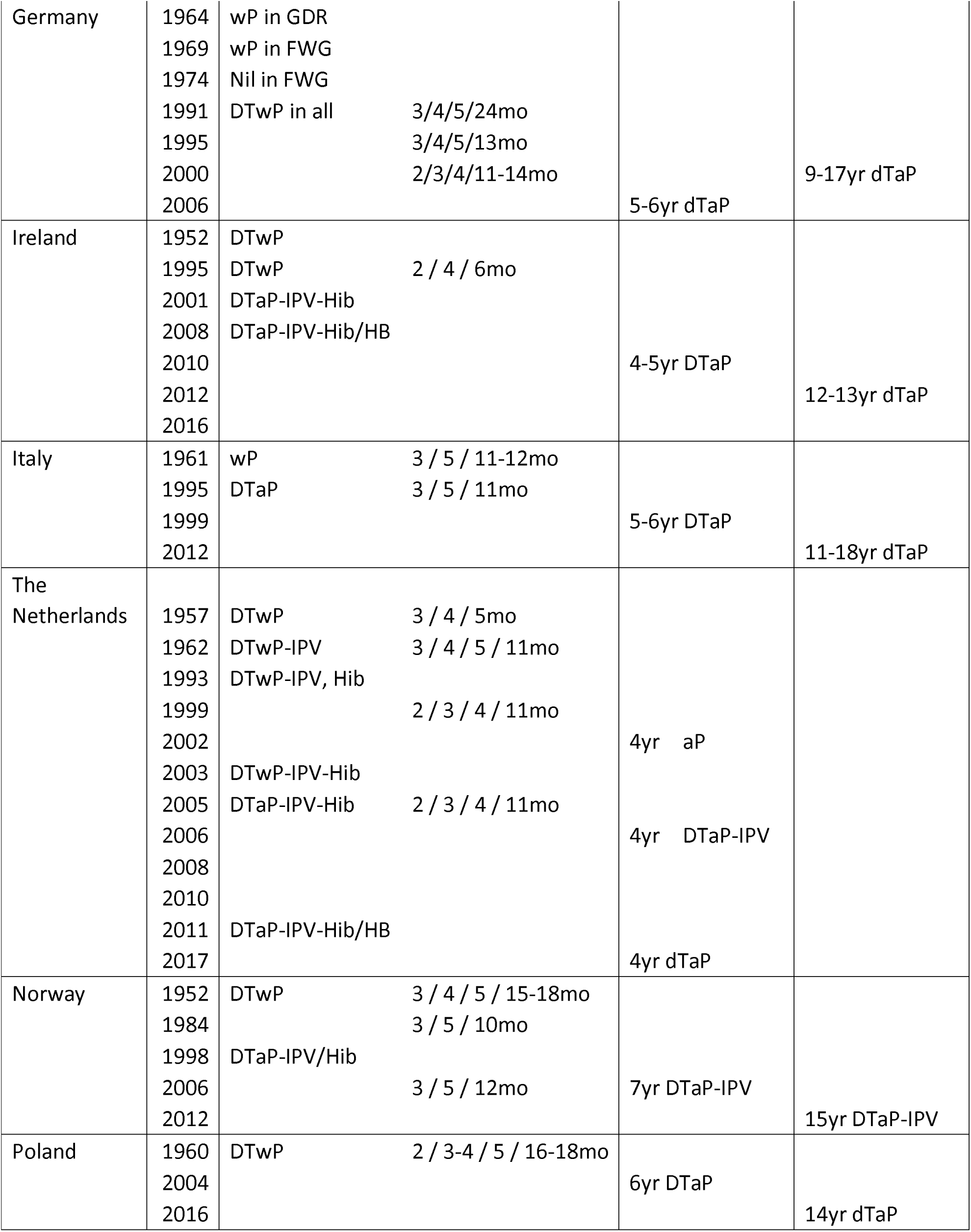
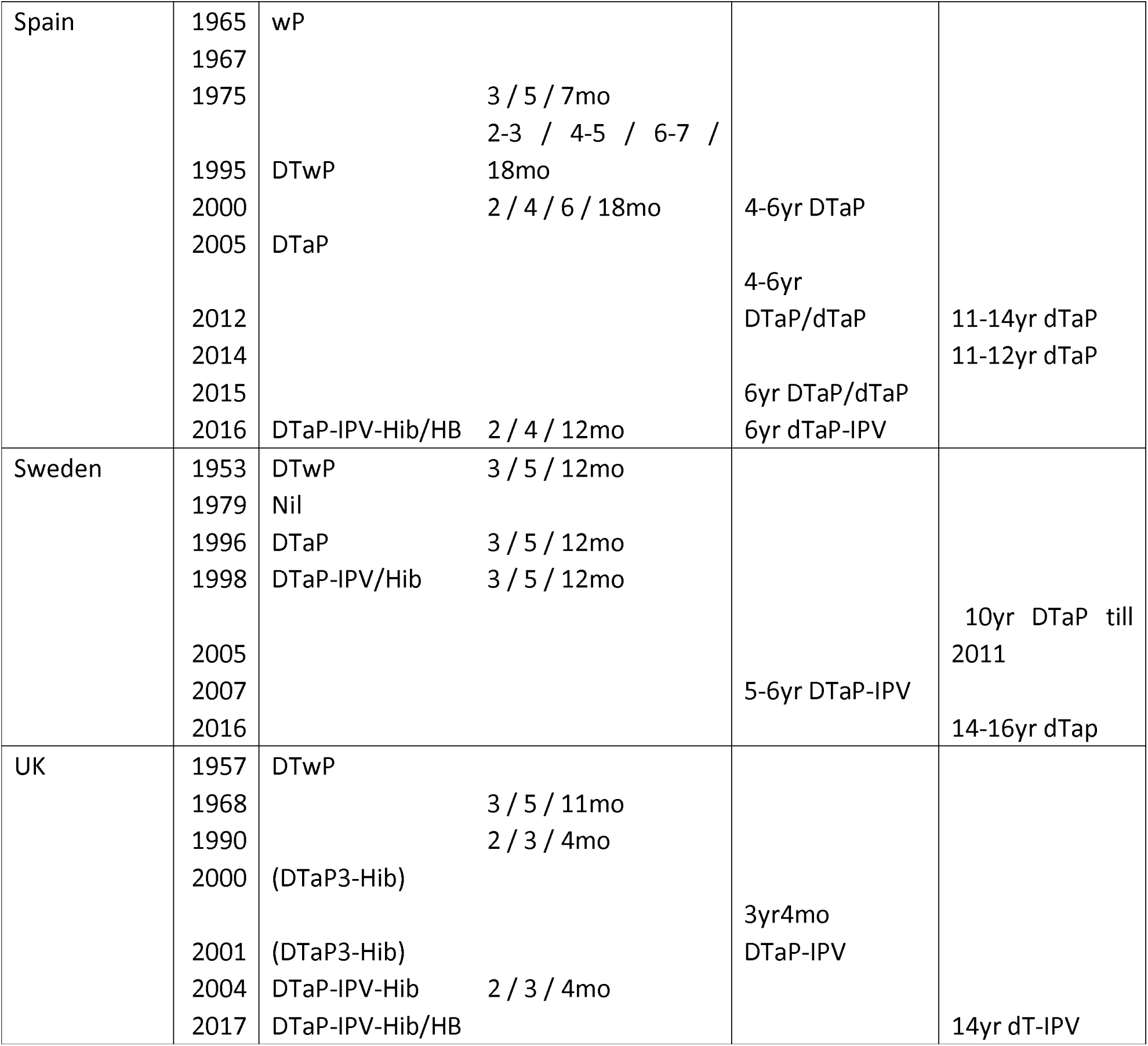

